# Sex-stratified autosomal association analysis reveals dimorphic genetic risk factors in non-systemic juvenile idiopathic arthritis

**DOI:** 10.64898/2026.09.08.26362497

**Authors:** Melissa Tordoff, Samantha L. Smith, Saskia Lawson-Tovey, UK JIA Biologics Register, CAPS, CHARMS, JIAGC, Lianne Kearsley-Fleet, Andrew D Smith, Stephanie J W. Shoop-Worrall, Michael W Beresford, Athimalaipet V. Ramanan, Stephen Eyre, Kimme L Hyrich, Lucy R Wedderburn, Andrew P Morris, John Bowes, CHYRRP and the CLUSTER consortium

## Abstract

**Objectives:** Juvenile idiopathic arthritis (JIA) is a collection of childhood onset rheumatic conditions that more frequently occur in females in most subtypes. Sex influences disease biology, treatment trajectories, and patient outcomes, however, no study to date has investigated the interaction of sex with autosomal genetic risk to JIA.

**Methods:** Genotype data from 3748 JIA cases (2551 female, 1197 male) and 9196 controls (5136 female, 4060 male) were analysed using a sex-stratified genome-wide association analysis. Systemic JIA was excluded. *HLA* imputation and fine-mapping were performed using SNP2HLA. Sexspecific effects were assessed using an omnibus sex-interaction test for multi-allelic amino acid positions. A subtype specific analysis was repeated in rheumatoid factor (RF) negative polyarthritis and oligoarthritis (1862 female, 718 male).

**Results:** A sex-combined analysis identified genome-wide significant (P<5x10^-8^) loci across the HLA region and nine loci outside the *HLA* region, including a novel association at rs231977 (*NUPR1)*. Markers within the *HLA* region were significantly sex-dimorphic (P<5x10^-8^) and nine loci outside of the *HLA* region were suggestive for sex dimorphism (P<5x10^-6^), including *SPRY2. HLA* fine-mapping identified lysine at position 70 of *HLA-B* conferring greater male JIA risk, and tyrosine at position 10 of *HLA-DRB1* conferring greater female JIA risk. Subtype specific analysis strengthened the association of tyrosine at position 10 of *DRB1* in females.

**Conclusions:** This study provides the first evidence that autosomal genetic risk factors contribute to JIA risk in a sex-dimorphic manner, mirroring known clinical observations, and highlighting the importance of incorporating sex-stratified methods in future JIA studies.

**Key Messages:** What is already known on this topic

- Sex dimorphism is well-established in non-systemic juvenile idiopathic arthritis (JIA) presentation, with females more frequently affected. Genetic studies have defined genetic susceptibility loci for JIA, however the contribution of genetic risk factors for sex dimorphism remains unknown.

What this study adds

- This is the first sex-stratified autosomal genetic study of JIA, revealing that markers within the HLA region are sex-dimorphic in their association to JIA susceptibility. Specifically, lysine at position 70 of *HLA-B* presented a greater risk to JIA in males. Tyrosine at position 10 of *HLA-DRB1* presented a greater risk to JIA in females.

How this study might affect research, practice or policy

- In defining autosomal genetic risk that significantly differs between males and females with non-systemic, this study advances understanding of the biological mechanisms underlying sex dimorphism in JIA presentation. These findings highlight the importance of incorporating sex-stratification into future JIA genetic studies and may inform future research of JIA classification. Further validation is required before clinical translation of these findings.

## Introduction

Juvenile idiopathic arthritis (JIA) defines a heterogenous group of chronic inflammatory rheumatic conditions with onset before the age of 16 and with symptoms lasting for at least six weeks (1). The International League of Associations for Rheumatology (ILAR) recognises seven JIA subtypes, which vary in clinical presentation from age of onset, joint involvement and extra-articular features (2). Despite advances in understanding the aetiology of disease, JIA presents a significant burden on patient quality of life including chronic pain and functional disability (3). Sex dimorphism across ILAR subtypes is well-documented in the presentation of JIA, it is typically more frequent in females than in males with an overall ratio of 2:1. However, this sex bias varies between ILAR subtypes. Thus in oligoarthritis and rheumatoid factor (RF) negative polyarthritis the ratio is reported as around 3:1 (F:M), systemic JIA is reported to occur equally within the sexes, while enthesitis-related arthritis (ERA) is more prevalent in males with a ratio of around 1:3 (F:M) (1,4,5). Research in rheumatic diseases such as JIA, systemic sclerosis (SSc) and systemic erythematosus lupus (SLE) suggests that sex influences not only disease onset but severity, treatment response and lived experience (6). Disease course can be more aggressive and long-term functional outcomes can be poorer in females (7). Additionally Tuomi et al., 2025 reported that female oligoarticular JIA patients are less likely to reach remission or have lower chance of early remission in comparison to their male counterparts (7,8). However, due to limited knowledge and recognition of sex-related differences in JIA, current guidelines do not incorporate sex-specific or personalised therapeutic approaches (7,9).

Sex dimorphism is a phenomenon that is documented across autoimmune diseases, with disease manifesting more frequently in females (10). Differences in the immune system, hormones, differences in gene regulation and sex-dependent environmental factors have been considered when investigating the mechanism of dimorphism in presentation of autoimmune diseases between the sexes (11). The influence of the hormones estrogen, progesterone and androgens such as testosterone on the inflammatory functions of immune cells demonstrate that hormones play an important role (10,11). One study demonstrated that hormones after puberty in females and the X chromosome interact to increase type 1 IFN response, increasing the likelihood of juvenile onset SLE in females around puberty (12). Additionally, estrogen in post puberty cisgender females and postmenopausal cisgender females undergoing hormone replacement therapy (HRT) were found to have increased levels of class-switched memory B cells, with estrogen impacting immune responses in those with an XX karyotype. Genes on the X chromosome have been reported to contribute to dimorphic traits however, more recent studies have demonstrated that autosomal genes may also influence traits differently in females and males (13). One study aimed to investigate the autosomal mechanisms behind sex dimorphism in SSc onset (Female to male ratio 8:1) and found one male-specific genetic locus (*BCL11A)* and an additional seven female-specific loci, which demonstrates the influence of autosomal loci on the sex bias of this disease (14).

Previous genetic studies have defined JIA susceptibility including risk markers within the Human leukocyte antigen (*HLA)* region and 22 genetic risk loci outside of the *HLA* region (15–17). However, the contribution of genetic risk to the difference in JIA onset between the sexes is unknown. Sex-specific genome-wide association studies (GWAS) have been developed to increase the statistical power to detect loci with different effects in males and females when compared to traditional sex-combined GWAS methods (18). Understanding of the mechanism behind sex dimorphism within JIA will advance understanding in the aetiology of the disease and contribute to advancements in diagnostics and therapeutic care to address a health disparity in outcome across sexes. Therefore, this study aimed to investigate genetic autosomal sex dimorphism in a cohort of JIA patients by implementing a sex-stratified GWAS.

## Methods

### Study Cohort

Patients were recruited by the following UK JIA cohort studies: the UK JIA Biologics Register, which includes the British Society for Paediatric and Adolescent Rheumatology Etanercept cohort study (BSPAR-ETN) and the Biologics for Children with Rheumatic Diseases study (BCRD) (19), the Childhood Arthritis Prospective Study (CAPS) (20), Childhood Arthritis Response to Medication Study (CHARMS) (21) and United Kingdom Juvenile Idiopathic Arthritis Genetics Consortium (UKJIAGC) (22). Genetic data for 9,196 population controls was available through the UK Household Longitudinal Study (23).

### Genotyping and Imputation

Genotyping data was available for 3748 cases of JIA, genotyped using the Illumina Infinium CoreExome array as described previously (15) (Supplementary table 1). Population controls were genotyped at the Wellcome Trust Sanger Institute using the Illumina Infinium CoreExome genotyping array. Quality control (QC) measures included exclusion of samples with a call rate <0.98, that had a discrepancy between genetically inferred sex and database records, that were identified by identity-by-descent (IBD) as related and were identified as ancestral outliers based on principal component analysis (PCA) using the flashpca software package (version 2.0). Individuals with inferred European ancestry were retained for analysis. Binary sex was defined by genotype cross-referenced with clinical records. Systemic JIA cases were excluded from this study as sex dimorphism is not reported in this ILAR subtype.

Additionally, individuals that were missing data for sex were excluded. Thus, a total of 3748 JIA cases were included in this study, 2551 female and 1197 male. Single nucleotide polymorphisms (SNPs) were excluded if they were non-autosomal, had a call-rate <0.98 and a minor allele frequency (MAF) <0.01. These quality control measures retained 7,684,338 highquality SNPs for analysis. Genome-wide imputation was performed using SHAPEIT5 (v5.1.1) for phasing and IMPUTE5 (v1.1.5) using the HRC reference panel (r1.1 EGAD00001002729).

Imputed variants with an information score <0.4 and MAF <1% were excluded (24,25). Two and four digit HLA alleles, amino acid residues and SNPs within the major histocompatibility complex (MHC) on chromosome 6 (29-34Mb hg build 19) were imputed using SNP2HLA software package (v1.0.3) (26). SNP2HLA utilises Beagle for phasing and imputation using the T1DG reference panel. The imputed MHC dataset was filtered for variants with an information score >0.9 and MAF >1% (26). A total of 7773 HLA markers remained for analysis.

### Genetic association analysis

A logistic regression was performed to generate summary statistics for variant associations in female and male datasets separately and for the combined dataset with the top three principal components as covariates and adjusted for sex using PLINK2 (v2.0.0) (27). Sex dimorphism analysis was performed using a sex-differentiated genome-wide meta-analysis (GWAMA) using the GWAMA software package (18,28). Using a sex-differentiated GWAMA generates a sex-differentiated p-value (pdiff) and a heterogeneity p-value (phet). The pdiff value represents the association of the genetic marker with JIA while allowing for differences in estimates between the sexes, thus representing a GWAS for JIA with greater power to detect an association in the presence of sex-dimorphic effects. The phet value tests the heterogeneity of a genetic marker’s association to JIA between the sexes thus giving a measure of sex dimorphism (18). A sex dimorphism analysis was performed on imputed HLA data by generating summary statistics for each HLA marker in males and females separately using a logistic regression and conducting a sex-differentiated GWAMA. An omnibus test was performed at each multi-allelic amino acid position using a log-likelihood ratio test of the null model and fitted model including all amino acid residues at the position using the most frequent residue as the reference category. Sex-specific effects of HLA amino acid positions were evaluated using an omnibus sex-interaction test by comparing a reduced model including the main effects of sex, amino acid residues, and genetic principal components to a full model, which additionally included sex-by-residue interaction terms for all non-reference residues. A subtype specific analysis was performed by restricting cases to oligoarthritis and RF negative polyarthritis, representing a more homogenous disease group (1862 female, 718 male) and repeating all analyses.

### Single-cell data and analysis

Single-cell RNA-sequencing count matrices and accompanying metadata were obtained from the published MAP-JAG JIA single-cell atlas (GEO accession: GSE278962). The downloaded count matrices had already undergone QC filtering as previously described (29). Analyses were performed in R version 4.3.3 using Seurat v5.3 (30). The expression assay was log-normalised using the *NormalizeData()* function, variable features were identified using *FindVariableFeatures()* and data were scaled using *ScaleData()* whilst regressing out cell-cycle scores and mitochondrial gene content. Samples were integrated using reciprocal PCA (RPCA)-based integration with 30 dimensions. Cell-type annotations were taken directly from the published atlas and used for all downstream analyses. Cell populations containing fewer than 50 cells were excluded from analysis.

The analysed dataset comprised 102,026 synovial tissue cells from 6 female and 3 male patients and 63,054 synovial fluid cells from 7 female and 3 male patients. Expression of candidate genes mapping to suggestive sex-dimorphic GWAS loci were examined across synovial fluid and tissue populations (Table 2). Genes lacking detectable expression within a lineage were excluded from lineage-specific visualisation. *BPNT2* was not detected in the dataset and *HTR1E* showed no detectable expression across synovial fluid or tissue populations and were excluded from downstream analyses. Additionally, B-cells showed no detectable expression of any sex-dimorphic genes and were thus omitted from the plots.

For heatmap generation, expression was aggregated at the sample and cell-type level using *AggregateExpression().* Aggregated expression values were averaged within sex for each annotated cell population. Expression matrices were separated by lineage and scaled independently prior to visualisation. Violin plots were generated from single-cell normalised expression values using the published atlas annotations. Heatmaps therefore represent average expression across all cells within a population, whereas violin plots provide single-cell resolution and allow assessment of the proportion of expressing cells within each population. Heatmaps were generated using the ComplexHeatmap package (v2.18) and violin plots were generated using Seurat using the *VlnPlot()* fucntion.

## Results

### Sex-combined JIA GWAS

A sex-combined genome-wide association analysis of the cohort identified markers reaching genome-wide significance (5x10^-08^) for JIA susceptibility across the *HLA* region and ten independent loci outside of the *HLA* region (Figure 1, Table 1). The top marker for the analysis was located within the *HLA* region, rs9272218 (*HLA-DQA1), SexCombinedP* = 4.31x10^-122^, further fine-mapping of the *HLA* region is described below. Five of the markers outside of the *HLA* region mapped to known susceptibility loci for JIA (15,17). The variant rs231977 mapping closely to *NUPR1* had not been previously reported in JIA susceptibility GWASs. When considering a suggestive significance threshold of 5x10^-06^, there were an additional 42 loci with suggestive association to JIA (Supplementary table 2). Restricting to the RF negative polyarthritis and oligoarthritis subtypes identified a similar set of associations across the *HLA* region with nine loci reaching genome-wide significance outside of the *HLA* region (Supplementary figure 1, Supplementary table 3).

**Figure 1.**
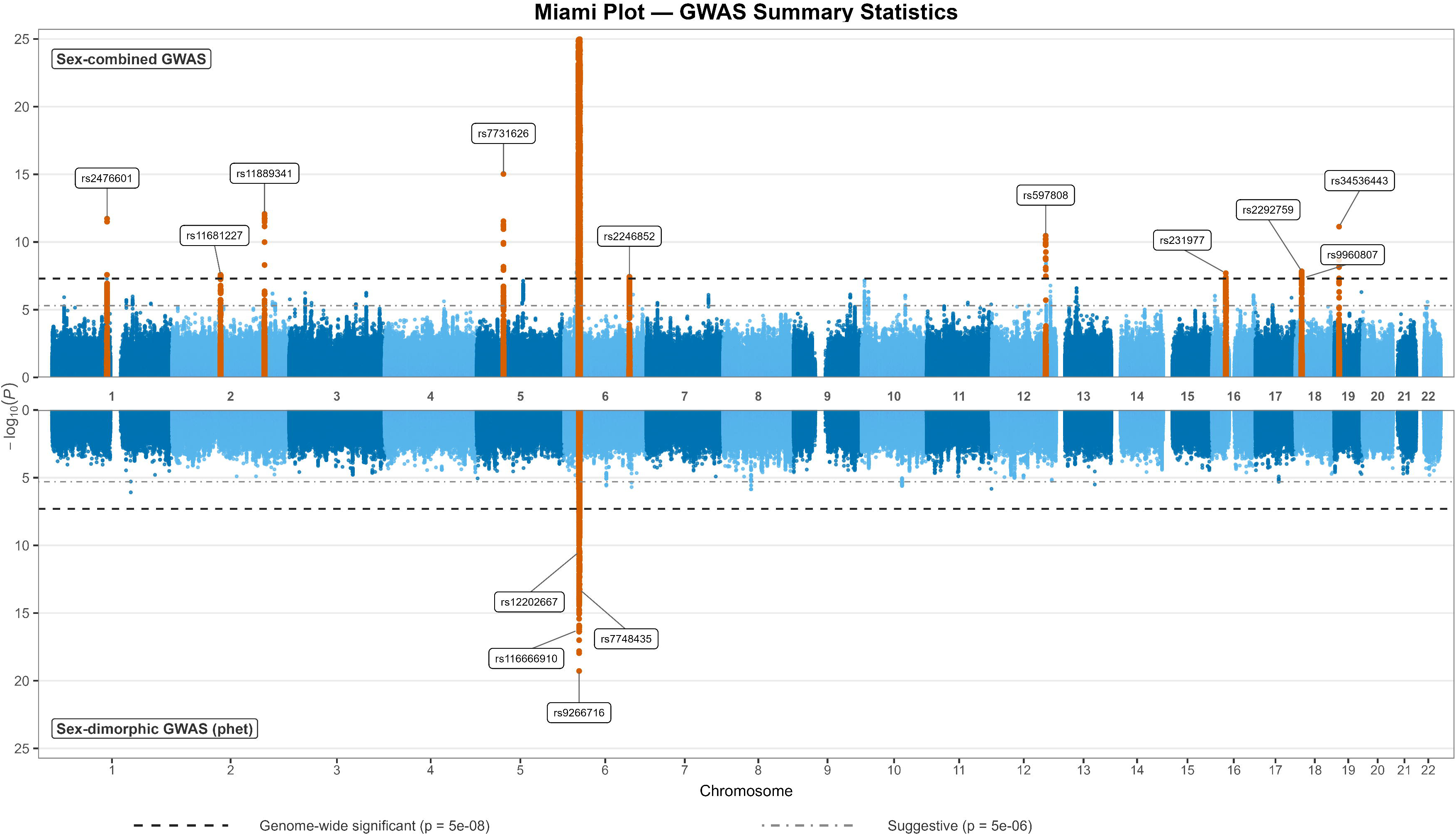
JIA GWAS associations and sex-dimorphic associations. Miami plot showing genome-wide associations of the sex-combined GWAS and sex-dimorphic associations of the sex-differentiated GWAMA. The -log10 of the p values are plotted against the chromosome position.

**Table 1.** Non-HLA loci that reached genome-wide significance in the sex-combined GWAS.

| SNP | CHR:BP | Notable Gene | Minor allele | MAF | Sexcombi<br>ned<br>P | Sexcombi<br>ned<br>OR (95CI) | pdiff | phet | Female P | Female OR<br>(95CI) | Male P | Male OR<br>(95CI) |
| --- | --- | --- | --- | --- | --- | --- | --- | --- | --- | --- | --- | --- |
| rs7731626 | 5:55444683 | ANKRD55 | A | 0.35 | 9.4x10-16 | 0.77 (0.72-0.82) | 1.0x10-14 | 0.99 | 1.1x10-10 | 0.77 (0.71-0.83) | 1.8x10-06 | 0.77 (0.69-0.86) |
| rs11889341 | 2:191943742 | STAT4 | T | 0.23 | 8.3x10-13 | 1.26 (1.18-1.34) | 5.7x10-12 | 0.041 | 1.1x10-11 | 1.31 (1.21-1.42) | 1.6x10-02 | 1.14 (1.02-1.27) |
| rs2476601 | 1:114377568 | PTPN22 | A | 0.10 | 1.8x10-12 | 1.36 (1.25-1.48) | 1.8x10-12 | 0.017 | 2.8x10-12 | 1.47 (1.32-1.64) | 2.2x10-02 | 1.18 (1.02-1.37) |
| rs34536443 | 19:10463118 | TYK2 | C | 0.04 | 7.3x10-12 | 0.56 (0.47-0.66) | 3.0x10-11 | 0.10 | 2.4x10-10 | 0.5 (0.41-0.62) | 3.9x10-03 | 0.67 (0.51-0.88) |
| rs597808 | 12:111973358 | ATXN2 | A | 0.49 | 3.4x10-11 | 1.21 (1.14-1.27) | 1.8x10-11 | 6.0x10-03 | 8.9x10-12 | 1.27 (1.19-1.37) | 9.4x10-02 | 1.08 (0.99-1.19) |
| rs2292759 | 18:12884343 | PTPN2 | A | 0.39 | 1.4x10-08 | 0.85 (0.80-0.90) | 1.9x10-07 | 0.88 | 4.9x10-06 | 0.85 (0.79-0.91) | 1.5x10-03 | 0.86 (0.78-0.94) |
| rs231977 | 16:28542172 | NUPR1 | T | 0.44 | 1.9x10-08 | 0.85 (0.80-0.90) | 1.1x10-06 | 0.78 | 1.2x10-05 | 0.85 (0.79-0.92) | 3.8x10-03 | 0.87 (0.79-0.96) |
| rs11681227 | 2:100754997 | AFF3 | G | 0.34 | 2.6x10-08 | 0.85 (0.80-0.90) | 8.7x10-07 | 0.69 | 6.1x10-05 | 0.86 (0.8-0.93) | 5.8x10-04 | 0.84 (0.76-0.93) |
| rs2246852 | 6:135691792 | AHI1 | G | 0.39 | 3.6x10-08 | 1.17 (1.10-1.23) | 1.4x10-09 | 6.5x10-04 | 2.1x10-10 | 1.25 (1.17-1.34) | 0.64 | 1.02 (0.93-1.12) |
| rs9960807 | 18:12770851 | LINC01882 | G | 0.13 | 4.9x10-08 | 1.24 (1.15-1.34) | 2.8x10-07 | 0.83 | 5.6x10-06 | 1.25 (1.14-1.38) | 2.1x10-03 | 1.23 (1.08-1.41) |

SNP, single nucleotide polymorphism; CHR, chromosome; BP, base position; MAF, minor allele frequency; OR, odds ratio; CI, confidence intervals; pdiff, sex-differentiated p-value; phet, heterogeneity p-value.

### Candidate loci for sex-dimorphic onset to JIA

A total of nine independent markers within the *HLA* region were significant for sex dimorphism for JIA susceptibility (*phet* <5x10^-08^) (Figure 1). The top marker for sex dimorphism (*phet*) was rs9266716 (*HLA-B), phet* = 5.23x10^-20^. Further *HLA* fine-mapping was necessary to define the genetic risk (see below). Eight loci outside of the *HLA* region were suggestive for significant (5x10^-6^) sex dimorphism for JIA susceptibility revealing opposite directions of effects in males and females, which would be masked by sex-combined analysis (Table 2, Supplementary figure 2). There was no evidence for significant sex-differentiated effects at the index SNPs identified in the sex-combined analysis. Within the RF negative polyarthritis and oligoarthritis sub-analysis, three markers within the *HLA* region reached genome-wide significance for sex dimorphism. Further fine-mapping of this region can be seen below. Outside of the *HLA* region, eight loci reached a suggestive threshold (5x10^-6^) for significance for sex dimorphism (Supplementary table 4).

**Table 2.**
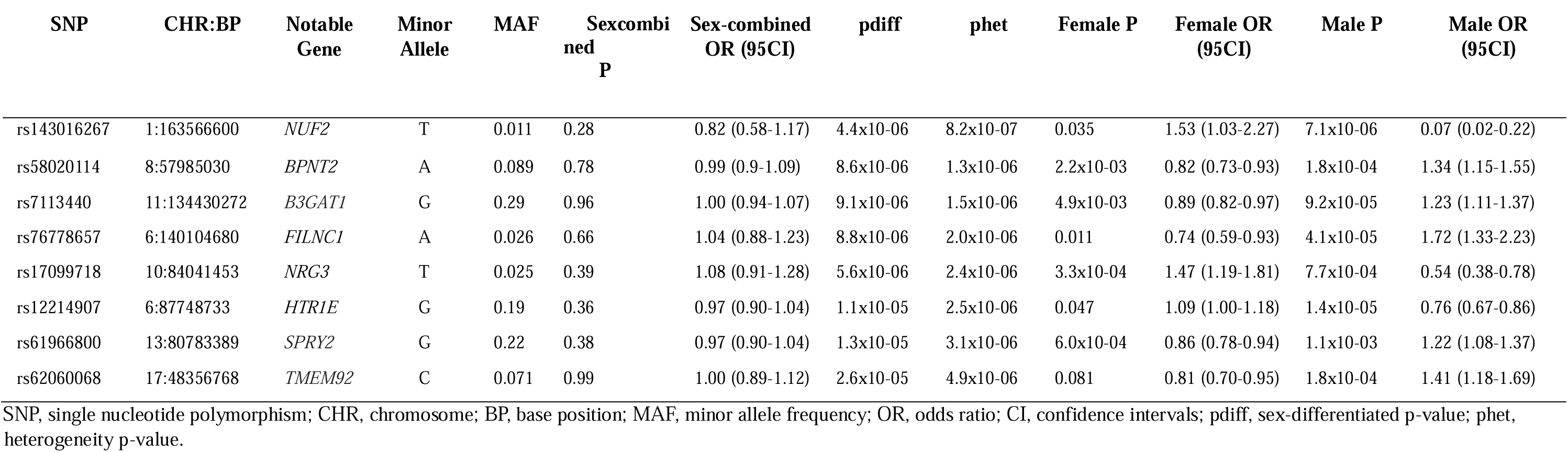
Non-HLA loci passing suggestive significance threshold (5x10-06) for sex dimorphism in a sex-differentiated GWAMA.

To identify the cellular populations through which sex-dimorphic genetic risk may act, expression of candidate genes mapping to suggestive sex-dimorphic GWAS loci were interrogated across synovial fluid and synovial tissue immune populations in the published MAP-JAG JIA single-cell atlas. Expression patterns varied substantially between genes with distinct patterns observed across lymphoid, myeloid and stromal populations (Figure 2). SPRY2 showed the broadest expression across both synovial fluid and synovial tissue cell populations, although expression was generally more prominent within synovial tissue. In contrast, NUF2 and NRG3 displayed highly restricted expression patterns, suggesting more specialised roles within specific cellular populations rather than broad effects across the synovial microenvironment. Similarly, FILNC1 and TMEM92 were expressed in only a limited number of populations, with FILNC1 largely confined to tissue myeloid cells and TMEM92 predominantly detected within stromal populations. B3GAT1 showed very low expression across the dataset, precluding meaningful interpretation. The highest proportion of SPRY2-positive cells was observed within stromal populations, while expression within Tcells was largely restricted to cytotoxic CD8+ granzyme positive populations. Expression tended to be higher in females, although differences were modest. Given its established role as a regulator of ERK/MAPK signalling pathways, SPRY2 may represent an additional candidate gene linking sex-associated genetic variation to immune and stromal cell activation.

**Figure 2.**
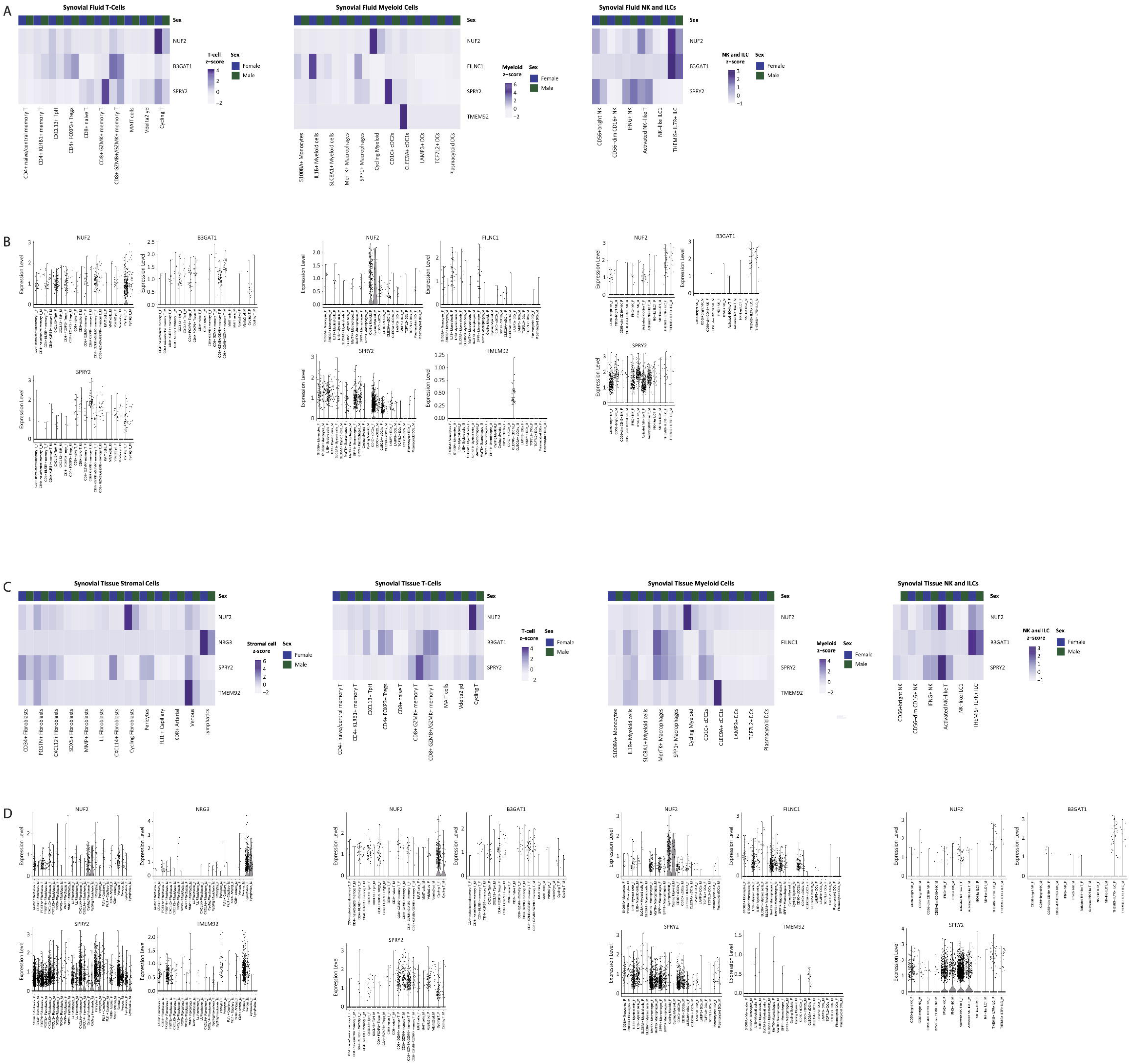
Expression of sex-dimorphic GWAS candidate genes across synovial fluid and tissue immune-cell populations. (A) Heatmaps showing aggregated expression of genes located within sex-dimorphic JIA GWAS loci across synovial fluid immune-cell populations. Expression was aggregated at the patient level, averaged by sex and scaled within lineage (T-cells, myeloid cells, NK/ILC and B cell populations). Rows represent genes and columns represent cell populations stratified by sex (female [F] and male [M]). Gene expression is colour-coded using average scaled expression values per population, based on a zscore distribution, ranging from low (white) to high expression (purple). (B) Violin plots showing single-cell expression of sex-dimorphic GWAS candidate genes across synovial fluid immune-cell populations. The x-axis shows cell populations stratified by sex and the y-axis shows normalised gene expression. Each violin represents the distribution of expression values across individual cells. (C) Same as (A) for synovial tissue immune cell populations (Stromal cells, T-cells, myeloid cells, NK/ILC and B cell populations). (D) Same as (B) for synovial tissue immune cell populations.

### Evidence of sex dimorphism for JIA susceptibility within the *HLA* region

The most significant association in the sex-combined analysis was to the previously reported amino acid position 13 of *HLA-DRB1* (*Sex-combined OmnibusP* = 5.46x10^-189^). Sexstratified fine-mapping of the *HLA* region confirmed that markers within *HLA-B* and *HLADRB1* presented the most significant associations to JIA susceptibility in a sex-dimorphic manner (Figure 3). A sex-stratified omnibus test of amino acid positions demonstrated that position 70 within *HLA-B, OmnibusP* = 5.03x10^-20^, and position 10 within *HLA-DRB1, OmnibusP* = 2.01x10^-11^, were dimorphic for JIA susceptibility. There was also significant association evidence to support a sex-dimporhic effect at the previously reported susceptibility association to amino acid position 11 and 13 of *HLA-DRB1* (*OmnibusP =* 1.96x10^-10^ and 7.04x10^-11^ respectively). Individual univariate analysis of each residue at the amino acid positions with sex as an interaction term revealed that lysine at position 70 of *HLA-B* presented the most risk in a sex-dimorphic manner to JIA (*SexInteractionP* = 2.6x10^-17^), with a greater effect size in males (*MaleOR* = 4.22, *FemaleOR* = 1.65). Tyrosine and glutamine at position 10 of *HLA-DRB1* were dimorphic for JIA susceptibility with similar magnitude. (*SexInteractionP* = 2.08x10^-11^ and *SexInteractionP* = 1.04x10^-11^, respectively), Table 3. *HLADRB1* position 10 is in strong linkage disequilibrium with positions 11, 12 and 13, therefore summary statistics for these positions are presented in Table 3.When conducting a sexdifferentiated GWAMA of the *HLA* region, the top alleles for sex dimorphism are *HLA-B*2705* (phet 3.22x10^-18^, *MaleOR* = 4.43, *FemaleOR* = 1.62) and *HLA-DRB1*08* (phet 6.39x10^-08^, *MaleOR* = 2.51, *FemaleOR* = 5.36). The analysis was repeated in the RF negative polyarticular and oligoarticular subtypes, where the association at *HLA-DRB1* position 10 remained genomewide significant *OmnibusP* = 3.82x10^-09^ and the association of position 70 of *HLA-B* more modestly associated at *OmnibusP =* 5.30x10^-06^, Supplementary figure 3 and Supplementary table 5.

**Figure 3.**
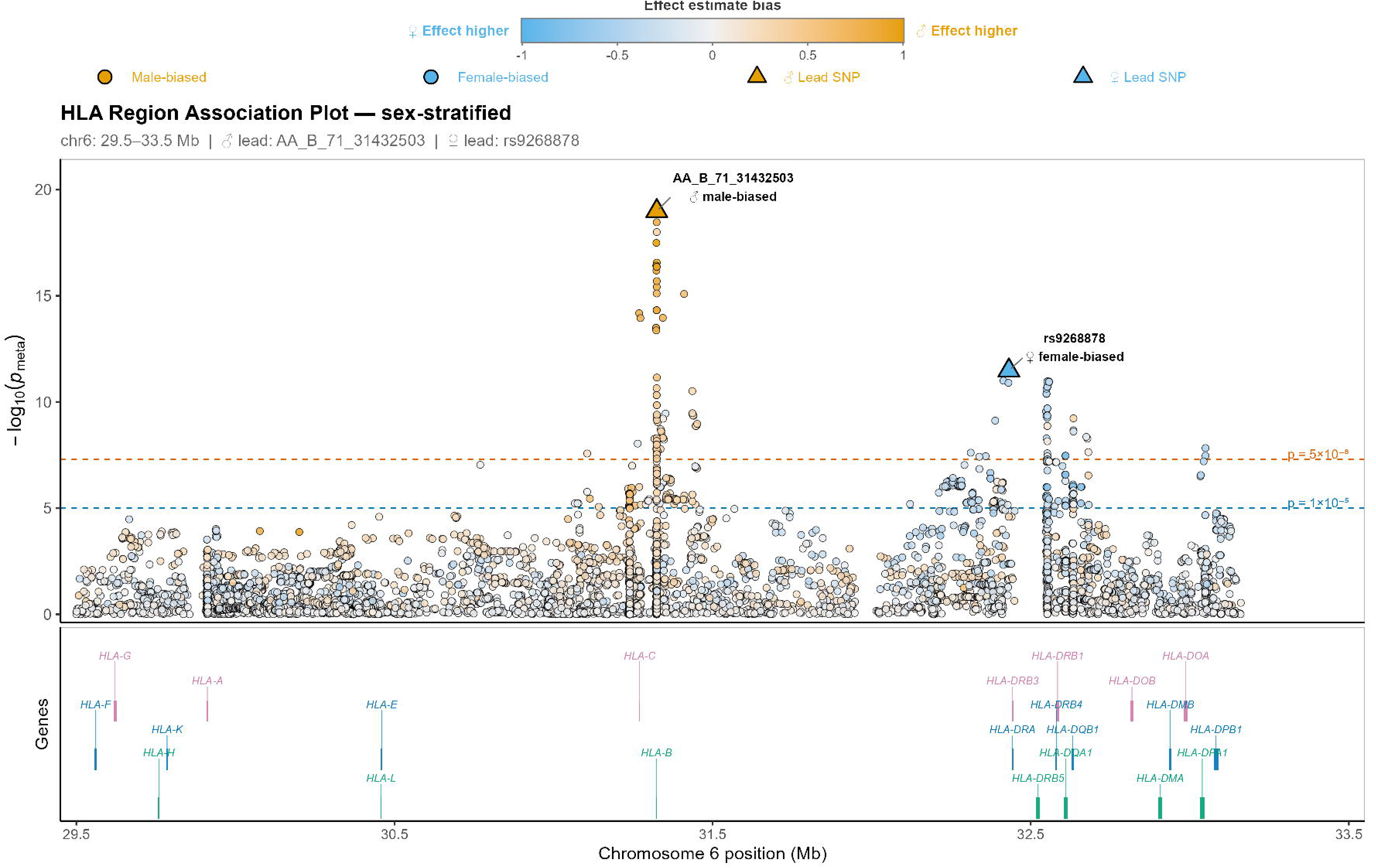
Sex-stratified association plot of the HLA region. Association results from a sex-stratified genome-wide association study are shown across the HLA region of chromosome 6 (GRCh37/hg19; 29.0–35.0 Mb). Each point represents a genetic variant, with genomic position on the x-axis and the meta-analysis −log (*p*-value), combining male and female summary statistics, on the y-axis. Points are coloured on a diverging scale reflecting the relative magnitude of the sex-specific effects: orange indicates variants where the risk is biased towards males, sky blue indicates variants where the risk is biased towards females, and intermediate colours indicate broadly similar effect sizes in both sexes.

**Table 3.**
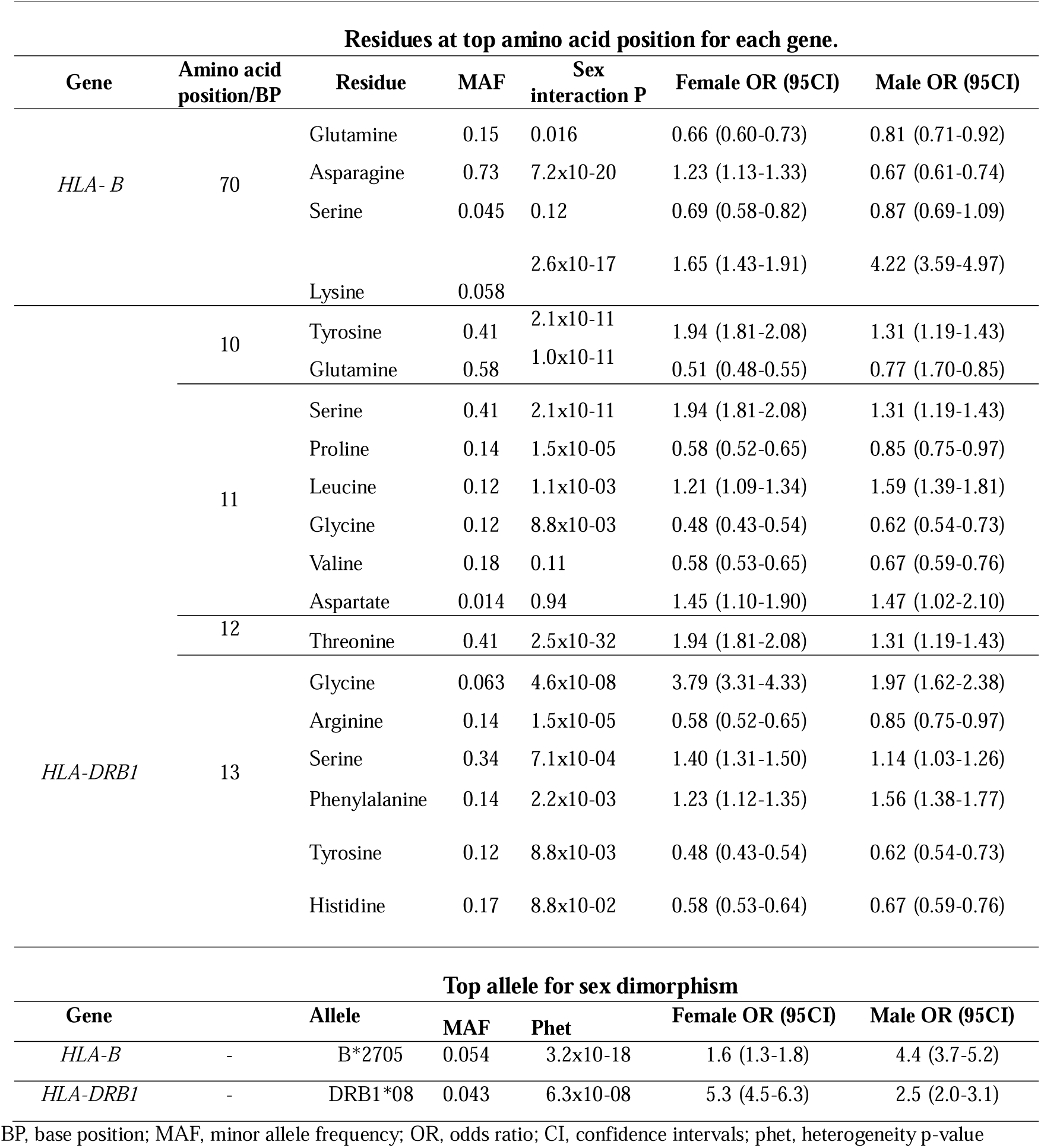
Associations and effect estimates for amino acids and alleles within sexdimorphic genes in the *HLA* region.

## Discussion

This is the first investigation of the sex-dimorphic effects of autosomal genetic risk factors for JIA susceptibility. Using a cohort of 3748 children and young people with nonsystemic JIA, we were able to detect a novel genome-wide association for JIA susceptibility: rs231977 with the closet gene being *NUPR1*. This variant is located on the locus 16p11.2, which is a known susceptibility locus for chronic inflammatory diseases. A recent JIA GWAS by López-Isac *et al.,* 2021 presented this locus as an “attractive biological candidate” for association with JIA susceptibility (15). This locus is closely located to *IL27*, which has been shown to be significantly elevated in blood plasma of JIA patients when compared to controls and reported as a risk marker for JIA in a recent proteome-wide association analysis (PWAS) (31,32). Within the RF-negative polyarthritis and oligoarthritis sub-analysis, rs2910686 within *ERAP2* gained strength in its association with JIA and reached genome-wide significance (*SexCombinedP* = 2.91×10), a stronger signal than seen in the whole cohort. *ERAP2* is a previously identified susceptibility locus for JIA (15). Similarly, in the subtype-restricted analysis, rs7977709 within *KDM2B* gained strength in its association with JIA susceptibility, reaching genome-wide significance and representing a novel JIA association (*SexCombinedP =* 1.10x10^-08^).

Using a sex-differentiated GWAMA, we demonstrated that markers within the *HLA* region are associated to JIA susceptibility with significant difference between females and males and eight non-HLA loci were suggestive for sex dimorphism for JIA. *SPRY2* (*phet =* 3.15x10^-06^) was among the most broadly expressed sex-dimorphic GWAS candidate genes in the singlecell data and was particularly enriched within synovial tissue stromal populations, including CD34+, CXCL12+ and CXCL14+ fibroblasts. Expression was generally higher in females, although the magnitude of these differences was modest. The stromal enrichment of *SPRY2* is notable given previous evidence demonstrating that *SPRY2* suppresses inflammatory responses in rheumatoid arthritis fibroblast-like synoviocytes through inhibition of ERK and AKT signalling pathways. Experimental overexpression of SPRY2 reduced cytokine production, matrix metalloproteinase expression and cellular proliferation in TNFα-stimulated synoviocytes, supporting a role as a negative regulator of synovial inflammation (33). The female-biased expression observed here may therefore reflect a compensatory regulatory mechanism acting to restrain inflammatory activation within the synovial microenvironment, although further functional studies will be required to determine the consequences of altered SPRY2 expression in JIA.

Fine-mapping of the *HLA* region detected that lysine at position 70 demonstrates a greater magnitude of risk of JIA in male patients (*MaleOR* = 4.22, *FemaleOR* = 1.65). Additionally, the association of the allele *HLA-B*2705* was sex-dimorphic for JIA susceptibility in this cohort, associated to increased risk of JIA in males (*MaleOR* = 4.43, *FemaleOR* = 1.62). *HLA-B*2705* is an established risk allele for ERA and ankylosing spondylitis (AS), which occurs more frequently in males (1,34). Positions 10, 11, 12 and 13 of *HLA-DRB1* and *HLA-DRB1*08* were associated with increased risk of JIA with greater magnitude in female patients. Tyrosine at position 10, serine at position 11 and threonine at position 12 make up the YST motif for the antigen binding grove of HLA-DRB1 and are in strong linkage disequilibrium with position 13 (35). Associations for tyrosine at position 10, serine of position 11 and threonine at position 12 of *HLA-DRB1* demonstrated significance for sex dimorphism and equal magnitude to JIA risk was detected among the amino acid positions for females in this cohort (*FemaleOR* = 1.94, *MaleOR* = 1.30). Glycine at position 13 of *HLADRB1* also demonstrated significance for sex dimorphism (*OmnibusP* = 7.04x10^-11^), with a higher JIA risk in female patients (*FemaleOR*= 3.78, *MaleOR*= 1.96). Studies of sex dimorphism have found that genetic markers within *HLA-DRB1* and *DQB1* have been reported to significantly vary in frequency between female and male type 1 diabetes patients, however, this is yet to be defined in JIA (36). Previous fine-mapping of the *HLA* region has reported associations of *HLA-DRB1*08* and positions 11 and 13 of *HLA-DRB1* for JIA susceptibility, where the cohort consisted of RF negative polyarticular arthritis and oligoarthritis subtypes (16). These JIA subtypes are more frequent in female patients (1). The allele *HLA-DRB1*08* was found to be more modestly dimorphic for JIA onset in this study (*phet* = 6.39x10^-08^), with a greater magnitude of risk in female patients (*FemaleOR* = 5.36, *MaleOR* = 2.51).

Repeating the analysis in RF negative polyarthritis and oligoarthritis subtypes demonstrated that *HLA-DRB1*08* remained associated to JIA susceptibility in a sex-dimorphic manner to a more modest magnitude (*phet =* 2.98x10^-05^), while *HLA-B*2705* lost strength in its association in JIA susceptibility in a sex-dimorphic manner (*phet =* 4.29x10^-04^). When repeating *HLA* fine-mapping in RF negative polyarthritis and oligoarthritis subtypes, position 70 of *HLA-B* remained modestly sex-dimorphic for JIA susceptibility (*OmnibusP* = 5.30x10^06^). Lysine at position 70 of *HLA-B* maintained a greater risk to JIA in males, however to a lesser magnitude when compared to the whole cohort (*MaleOR*= 2.34, *FemaleOR*= 1.47). Position 10 of *HLA-DRB1* remained dimorphic for JIA susceptibility (*OmnibusP*=2.82x20^-09^), with the magnitude of risk to JIA in females increased when compared to the whole cohort analysis (*FemaleOR* = 2.28, *MaleOR* = 1.50).

Given the well-established sex dimorphism in JIA presentation, this study utilised a sex-stratified genome-wide association study to provide power to detect genetic associations that may vary in direction of effect between the sexes and markers that are sex-dimorphic for JIA onset. Therefore, a strength of this study is the utilisation of this method in investigating genetic associations to JIA onset. However, the number of male JIA cases were fewer in this study in comparison to female JIA cases. This accurately reflects the ratio of JIA seen clinically, however the statistical power to detect associations is lowered by the imbalance in sample size between sexes (1). Furthermore, variants were not functionally validated in this study and interpretation should take this into consideration. Future research should include sex in functionally validating candidate regions for JIA onset. Several limitations should also be considered when interpreting the single-cell findings. Candidate genes were selected based on proximity to sex-dimorphic GWAS signals; however, the nearest gene is not necessarily the causal gene mediating disease risk. Many autoimmune risk variants reside within regulatory regions and may influence the expression of distal genes through long-range chromatin interactions. The expression patterns reported here should be interpreted as supporting the biological relevance of these loci, rather than identifying causal genes. In addition, a relatively small number of male samples were available within the published JIA single-cell dataset. Observed sex differences should therefore be considered exploratory. A major strength of the dataset, however, is the inclusion of both synovial fluid and synovial tissue samples, enabling assessment of gene expression across multiple disease-relevant cellular compartments and providing insight into both immune and stromal contributions to JIA pathogenesis.

Research on sex differences across rheumatic disease has primarily focussed on sex chromosomes, hormones and immune cell composition. Recently, autosomal genetic contribution has begun to be explored in SSc (14). For the first time, this study aimed to investigate genetic autosomal sex dimorphism in children and young people with JIA. Using statistical methods to explore sex interaction in genetic risk to JIA, this study detected a novel association for JIA susceptibility when using a sex-combined GWAS, *NUPR1.* In addition to identifying eight suggestive non-HLA loci for sex dimorphism to JIA risk, lysine at amino acid position 70 of *HLA-B* was found to contribute to a greater risk to JIA in males and tyrosine at position 10 of *HLA-DRB1* was found to contribute to a greater risk to JIA in females. Known JIA susceptibility alleles *HLA-B*2705* and *HLA-DRB1*08* were also found to be sexdimorphic in their risk to JIA. The difference in *HLA* risk to JIA between the sexes observed in this study reflect what is seen clinically in the presentation of JIA ILAR subtypes. *HLAB*27* is known to be associated with ERA and to AS, which is more prevalent in males (34,37).

In addition, previous fine-mapping of the *HLA* region for JIA detected *HLA-DRB1* positions 11 and 13 as risk markers in a cohort of RF negative polyarthritis and oligoarthritis (16). This study demonstrates that the associations of positions 11 and 13 of *HLA-DRB1* are sexdimorphic for JIA risk, with a greater risk detected in females. This risk increases in magnitude when the cohort was restricted to RF negative polyarthritis and oligoarthritis subtypes, which are more prevalent in females (1). Thus, providing evidence that the *HLA* region has distinct contributions to JIA between the sexes with markers in *HLA-B* presenting greater risk to JIA in males and makers within *HLA-DRB1* presenting greater risk to females.

Given the emerging literature defining the difference in disease burden JIA presents between the sexes, it is important to research the biological mechanisms behind the difference in JIA presentation between males and females. By understanding this difference in genetic risk to JIA, we may be able to begin to explain the biological mechanism behind the sex dimorphism observed. This may facilitate a more tailored approach to genetic diagnosis in the future and may even offer personalised treatment options between the sexes. This study therefore highlights the importance for future genetic research to include sex-stratified analyses so that novel insights into sex-specific genetic risk can be defined. Ultimately, sex-dimorphic genetic markers identified in this study have the potential to be integrated into future sexspecific diagnostic testing, classification criteria and therapy options for JIA. However, further research is required to validate these findings.

## Data Availability

The data underlying this article cannot be shared publicly due to ethics. The data will be shared on reasonable request to the corresponding author.

## Acknowledgments and affiliations

CLUSTER is supported by grants from the Medical Research Council (MRC) [MR/R013926/1] and Versus Arthritis [Grant: 22084], Great Ormond Street Hospital Children’s Charity [VS0518], and Olivia’s Vision. This work is supported by the NIHR GOSH BRC, the NIHR Biomedical Research Centre: Manchester, the NIHR GOSH Biomedical Research Centre and the British Society for Rheumatology (BSR), and the “UK’s Experimental Arthritis Treatment Centre for Children, supported by Versus Arthritis (grant: 20621)”. The views expressed are those of the author(s) and not necessarily those of the NHS, the NIHR or the Department of Health. Wedderburn is additionally supported by Versus Arthritis (grant: 21593) at the Centre for Adolescent Rheumatology Versus Arthritis. Hyrich is additionally supported by the Centre for Epidemiology Versus Arthritis (grant: 21755) and the Centre for Genetics and Genomics Versus Arthritis (grant: 21754) at the University of Manchester, UK.

LKF is supported by Arthritis UK (23126). SSW is supported by the MRC [MR/W027151/1].

This study acknowledges the use of the following UK JIA cohort collections: The Biologics for Children with Rheumatic Diseases (BCRD) study (funded by Arthritis Research UK grant: 20747); The British Society for Paediatric and Adolescent Rheumatology Etanercept Cohort Study (BSPAR-ETN) (funded by a research grant from the British Society for Rheumatology (BSR); BSR has previously also received restricted income from Pfizer to fund this project; Childhood Arthritis Prospective Study (CAPS) (funded by Versus Arthritis UK, grant: 20542); Childhood Arthritis Response to Medication Study (CHARMS) (funded by Sparks UK, reference 08ICH09; and the Medical Research Council, reference MR/M004600/1), United Kingdom Juvenile Idiopathic Arthritis Genetics Consortium (UKJIAGC). This study also acknowledges the use of the following two UK-wide JIA-associated uveitis clinical trials: the SYCAMORE Trial (funded by Arthritis Research UK, grant: 19612 and the National Institute of Health Research Health Technology Assessment, grant: 09/51/01); and the APTITUDE Trial (funded by Arthritis Research UK, grant: 20659). Understanding Society: The UK Household Longitudinal Study is led by the Institute for Social and Economic Research at the University of Essex and funded by the Economic and Social Research Council. The survey was conducted by NatCen and the genome-wide scan data were analysed and deposited by the Wellcome Trust Sanger Institute. Information on how to access the data can be found on the Understanding Society website https://www.understandingsociety.ac.uk/.

## Members of the CLUSTER Consortium are as follows

Prof Lucy R. Wedderburn, Ms Zoe Wanstall, Ms Vasiliki Alexiou, Mr Fatjon Dekaj, Ms Bethany R Jebson, Dr Melissa Kartawinata, Ms Aline Kimonyo, Ms Eileen Hahn, Ms Genevieve Gottschalk, Ms Freya Luling Feilding, Ms Alyssia McNeece, Ms Fatema Merali, Ms Elizabeth Ralph, Ms Emily Robinson, Ms Emma Sumner (UCL GOS Institute of Child Health, London); Prof Andrew Dick, (UCL Institute of Ophthalmology, London); Prof Michael W. Beresford, Dr Emil Carlsson, Dr Joanna Fairlie, Dr Jenna F. Gritzfeld, Dr Oliver McClurg, Dr Karen Rafferty (University of Liverpool); Prof Athimalaipet V Ramanan, Ms Teresa Duerr (University Hospitals Bristol and Weston NHS Foundation Trust); Prof Michael Barnes, Ms Sandra Ng, (Queen Mary University, London); Prof Kimme Hyrich, Prof Stephen Eyre, Prof Soumya Raychaudhuri, Prof Wendy Thomson, Dr John Bowes, Ms Jeronee Jennycloss, Ms Saskia Lawson-Tovey, Dr Paul Martin, Prof Andrew Morris, Dr Stephanie Shoop-Worrall, Dr Samantha Smith, Dr Michael Stadler, Dr Damian Tarasek, Dr Melissa Tordoff, Dr Annie Yarwood (University of Manchester); Dr Chris Wallace, Dr Wei-Yu Lin (University of Cambridge); Prof Nophar Geifman (University of Surrey); Dr Sarah Clarke (School of Population Health sciences and MRC Integrative Epidemiology Unit, University of Bristol). Dr Thierry Sornasse (AbbVie Inc.) Dr Robert J Benschop, Dr Rona Wang (Eli Lilly) Daniela Dastros-Pitei MD, PhD, Sumanta Mukherjee, PhD (GlaxoSmithKline Research and Development Limited.) Dr Michael McLean, Dr Anna Barkaway (Pfizer) Dr Peyman Adjamian (Swedish Orphan Biovitrum AB (publ) (Sobi)) Helen Neale (UCB Biopharma SRL.) The CLUSTER Champions.

We are grateful to the CLUSTER champions and members of the CLUSTER consortium, who have provided valuable feedback for this work and for the future dissemination of this work to patients and parents. The authors would like to acknowledge the assistance given by Research IT and the use of the Computational Shared Facility at The University of Manchester. Ethical approvals were gained from the Northwest Greater Manchester Central Research Ethics Committee (BCRD), West Midlands Multicentre Research Ethics Committee (BSPAR-ETN), Northwest Multicentre Ethics Committee (CAPS: REC/02/8/104, IRAS 184042) and the Bloomsbury/Central London Research Ethics Committee (CHARMS: REC 05/Q0508/95, IRAS 172219). No additional ethical permissions were required for this analysis. Written informed consent was provided by guardians of participants and age-appropriate consent/assent was provided by participants themselves, where appropriate. AVR has received consulting fees and honorariums from AbbVie, Eli Lilly,

UCB, Astra Zeneca, Pfizer, Roche, Novartis, Alimera and SOBI, KH has received grants from Pfizer and BMS unrelated to work in JIA and honorariums from AbbVie and is an Associate Editor at ARD, LRW has received research grants from MRC, Versus Arthritis, AbbVie, SOBI, UCB, GSK and Great Ormond Street Children’s Charity and honorariums from Pfizer and Advanced Targeted Therapies. The data underlying this article cannot be shared publicly due to ethics. The data will be shared on reasonable request to the corresponding author.

