## Supplementary materials for "Sex-stratified autosomal association analysis reveals dimorphic genetic risk factors in non-systemic juvenile idiopathic arthritis"

^1^Centre for Genetics and Genomics Versus Arthritis, Centre for Musculoskeletal Research, Manchester Academic Health Science Centre, The University of Manchester, Manchester, UK. ^2^National Institute of Health Research Biomedical Research Centre: Manchester, Manchester Academic Health Science Centre, Manchester University NHS Foundation Trust, Manchester, UK. ^3^Arthritis UK Centre for Epidemiology, Centre for Musculoskeletal Research, Manchester Academic Health Science Centre, The University of Manchester, Manchester, UK. ^4^Department of Women’s and Children’s Health, Institute of Life Course and Medical Sciences, University of Liverpool, Liverpool L14 5AB, UK. ^5^Department of Rheumatology, Alder Hey Children’s NHS Foundation Trust Hospital, Liverpool L14 5AB, UK. ^6^Bristol Royal Hospital for Children, Bristol, UK. ^7^Translational Health Sciences, University of Bristol, Bristol, UK. ^8^Infection, Immunity and Inflammation Research and Teaching Department, UCL Great Ormond Street Institute of Child Health, London, UK. ^9^Centre for Adolescent Rheumatology at UCL, UCL Hospital and Great Ormond Street Hospital, London, UK. ^10^NIHR Biomedical Research Centre at Great Ormond Street Hospital, London, UK.

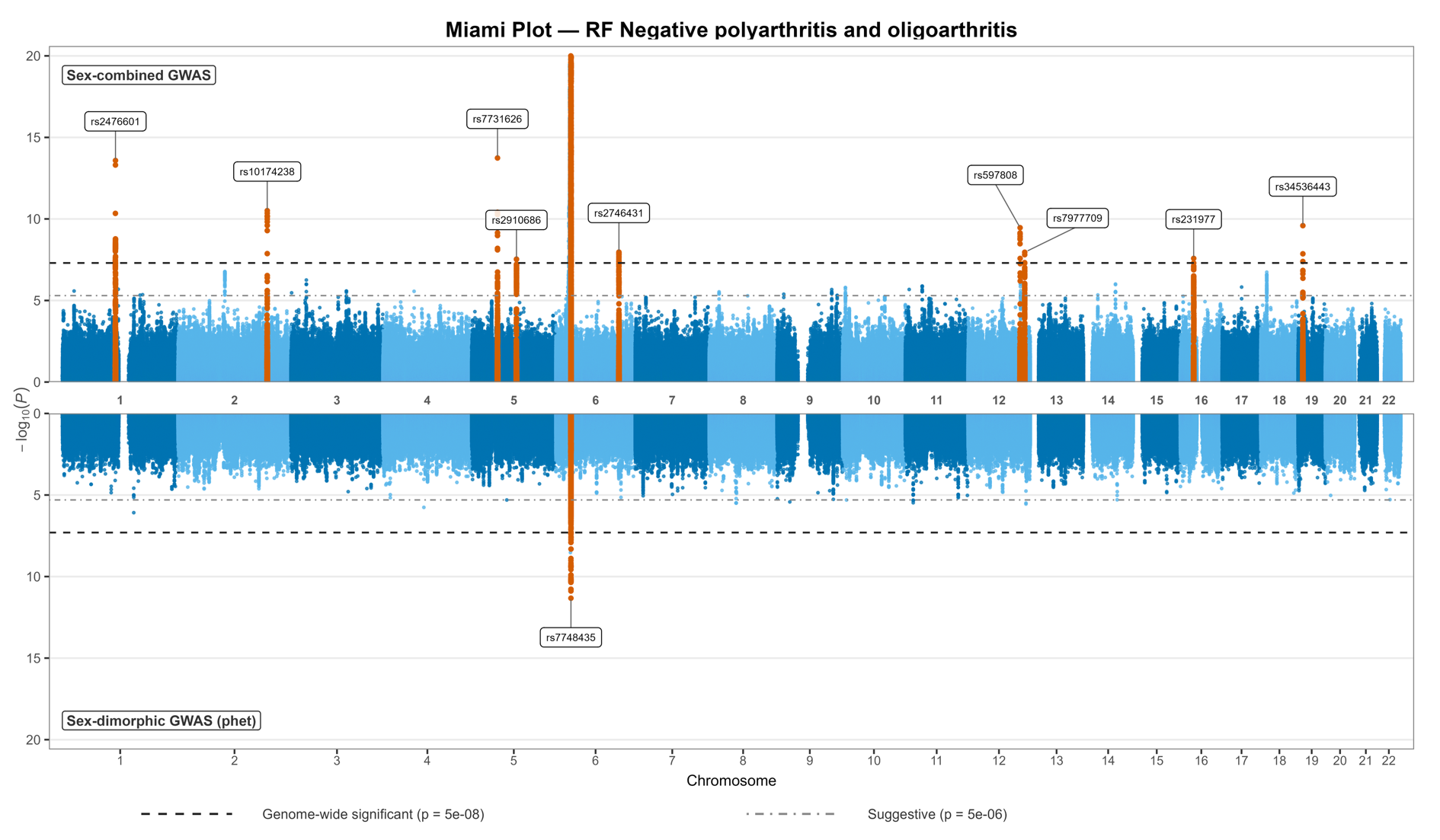
**Supplementary Figure 1 JIA GWAS associations and sex dimorphic associations of the RF negative polyarthritis and oligoarthritis.**

Miami plot showing genome-wide associations of the sex-combined GWAS and sex dimorphic associations of the sex-differentiated GWAMA. The -log10 of the p values are plotted against the chromosome position.

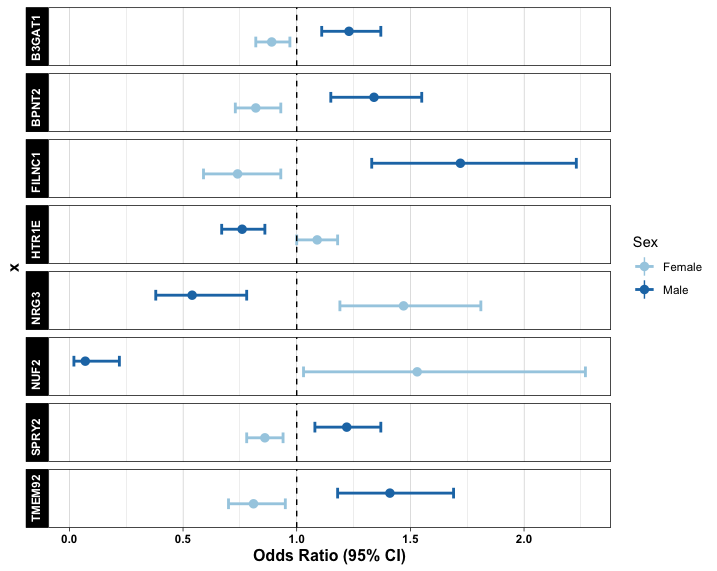
**Supplementary Figure 2 Effect sizes of males and females at nine non-HLA loci suggestive for sex dimorphism in non-systemic JIA**

Nine non-HLA loci reached suggestive significance (phet = 5x10^-06^) for sex dimorphism. Odds ratios are plotted with error bars representing 95% confidence intervals (CI). Black dashed line represents OR = 1.

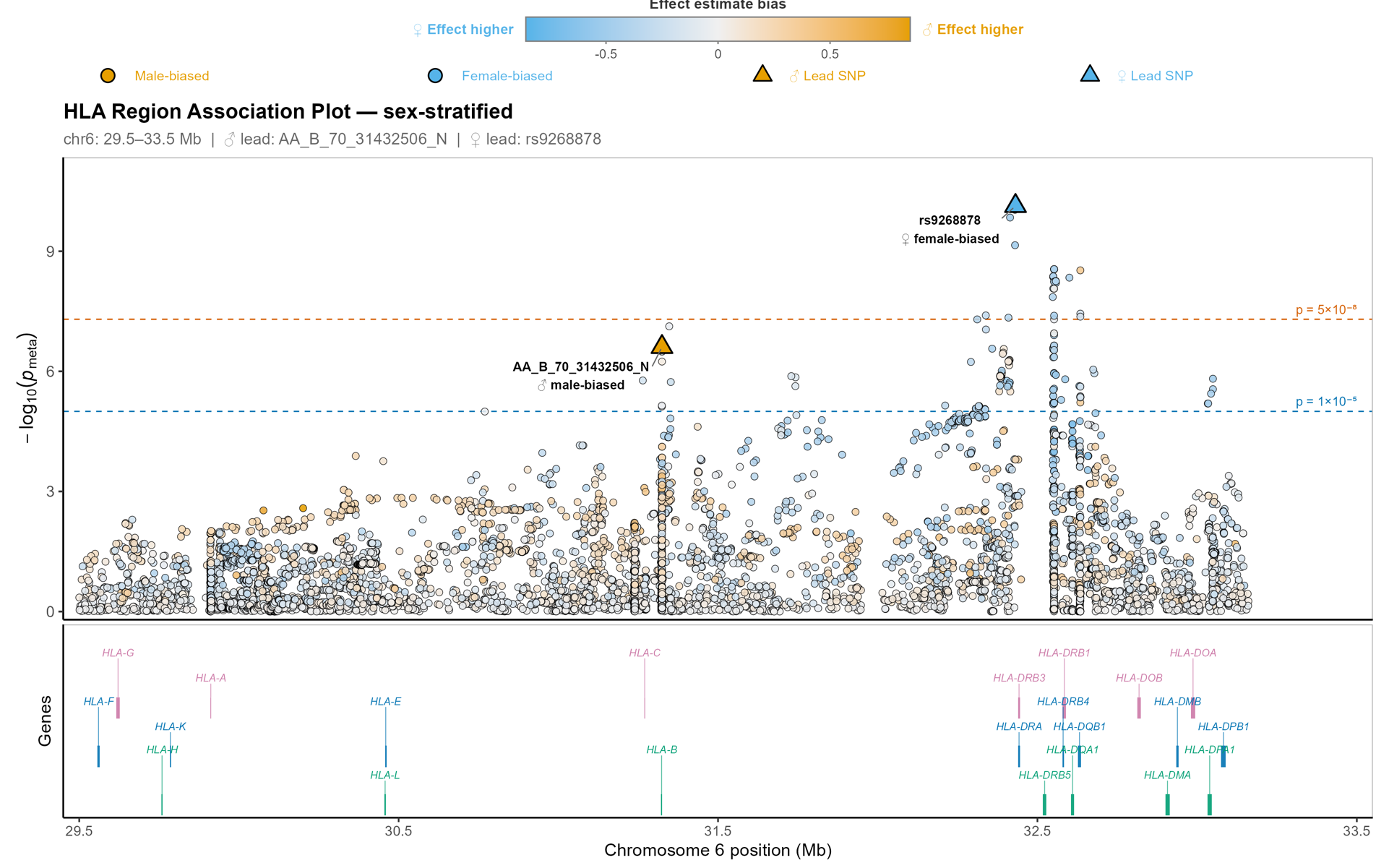
**Supplementary Figure 3 Sex dimorphic genetic markers of the *HLA* region for JIA susceptibility in rheumatoid factor negative polyarthritis and oligoarthritis.**

Manhattan plot of the *HLA* markers significant for sex dimorphism in the association to JIA onset in rheumatoid factor negative polyarthritis and oligoarthritis subtypes. Genome-wide significance (P<5x10^-08^) is indicated by the red dashed line. Suggestive significance (P<5x10^-06^) is indicated by the blue dashed line. The -log10 of the phet values are plotted against the position on chromosome 6. Blue points indicate a higher effect estimate in females, and yellow points indicate a higher effect estimate in males.
